# Growth patterns and ratios of posterior cranial fossa structures in the Japanese pediatric population: A study utilizing CT scans

**DOI:** 10.1101/2023.07.07.23292362

**Authors:** Hiroaki Hashimoto, Osamu Takemoto, Yasuyoshi Chiba

**Affiliations:** Department of Neurosurgery, Osaka Women’s and Children’s Hospital, Izumi, Osaka 594–1101, Japan; Department of Neurological Diagnosis and Restoration, Graduate School of Medicine, Osaka University, Suita, Osaka 565–0871, Japan

**Keywords:** brainstem volume, cerebellum volume, DICOM images, normal growth curves, pediatric patients, posterior cranial fossa volume

## Abstract

**Purpose:** The changes in the proportion of posterior cranial fossa structures during pediatric development remain unclear. This retrospective study aimed to investigate the growth patterns and ratios of these structures using computed tomography (CT) scans.

**Methods:** Head CT scans of pediatric patients with minor head trauma from our institute between March 2006 and May 2023 were analyzed. The study segmented the intracranial volume (ICV), posterior cranial fossa volume (PCFV), cerebellum volume (CBMV), and brainstem volume (BSV). Correlation coefficients were calculated among the parameters. Patients aged 0 to 10 years were divided into 15 age-related clusters, and mean and standard deviation values were measured. Growth curves were created by plotting mean values sequentially. Ratios such as PCFV/ICV and (CBMV + BSV)/PCFV were examined. Statistical analyses, including unpaired T-tests and logarithmic curve fitting, were performed.

**Results:** A total of 234 CT scans (97 from females, 115 from infants under one year of age) were analyzed. Positive correlations were observed among the parameters, with the strongest between PCFV and CBMV. The growth curves for ICV, PCFV, CBMV, and BSV exhibited a two-phase process, with rapid growth until approximately 4 years of age, followed by stabilization. The ratios PCFV/ICV and (CBMV + BSV)/PCFV showed increasing trends from birth onwards, stabilizing by 4 and one years of age, respectively.

**Conclusion:** This study provides insights into the growth patterns and ratios of posterior cranial fossa structures in the pediatric population. The findings demonstrate a two-phase growth process and increasing trends in the examined ratios.

## Introduction

Previous studies have documented a significant reduction in the ratio between the posterior cranial fossa volume (PCFV) and the supratentorial or intracranial volume in Chiari malformation type I [1, 2]. However, the association between the smaller volume of the posterior cranial fossa and Chiari malformation type I lacks conclusive statistical evidence [3–5]. In Chiari malformation type II, which is commonly observed in patients with myelomeningoceles (MMC), a small posterior fossa is recognized as one of the central nervous system malformations associated with Chiari malformation [6]. While cranio-cervical decompression has been performed in symptomatic cases of Chiari malformation type II [7], the mechanisms underlying symptom development, as well as the optional timing and surgical techniques, have not been fully elucidated [8]. Although a more rostral lesion level of MMC has been linked to a higher likelihood of requiring decompression surgery [9], the relationship between PCFV in Chiari malformation type II and the necessity for surgical decompression lacks statistical evidence.

Previous studies have assessed PCFV or cerebellum volume (CBMV) in relation to pediatric Chiari malformation type I [10] or type II [11], but they did not consider the influence of the normal growth of PCFV or CBMV. Considering the growth of the head and intracranial structures is crucial for the accurate assessment of Chiari malformation type I or type II in children. Therefore, investigating the normal growth pattern of PCFV or CBMV has the potential to provide further insights into the intricate relationship between Chiari malformation and the volume of posterior cranial fossa structures.

The objective of this study is to establish the normal growth pattern of posterior cranial fossa structures in the pediatric population. Previous studies have reported normal growth curves for PCFV, indicating two distinct phases: an initial phase characterized by rapid growth from birth to approximately 3-4 years of age, followed by a stabilization phase [12, 13]. However, the existing literature has certain limitations. Firstly, there is a lack of PCFV measurements in children under 3 months of age [12]. Additionally, the numbers of evaluated children under one year of age is limited, with only 20 patients included in one study [12]. Furthermore, while one study reported PCFV values at yearly intervals ranging from one to 16 years of age, detailed changes in PCFV during the first year of life have not been reported [13], despite the understanding that intracranial volume doubles by 9 months of age after birth [14]. Moreover, the changes in the proportion of posterior cranial fossa structures during pediatric development remain unknown. Therefore, this study aims to provide update and comprehensive data on the normal growth patterns and ratios of posterior cranial fossa structures to address these gaps in the existing literature.

## Material and methods

### Patients and study setting

We conducted a retrospective study involving patients who underwent computed tomography (CT) imaging for head trauma at Osaka Women’s and Children’s Hospital between March 2006 and May 2023. Considering the dose-response relationship between pediatric CT-related radiation exposure and brain cancer [15], our institution adheres to guidelines to determine the necessity of CT scans for children with head trauma [16, 17]. We excluded patients based on the following criteria: (1) suspicion of abuse, (2) requirement for craniotomy for decompression within a few days following head injury, (3) cranial depressed fracture necessitating reconstructive surgery, (4) evident compression and significant deformation of lateral ventricles due to hematoma, (5) Ommaya reservoir implantation or drainage surgeries, and (6) presence of complications such as craniosynostosis, tumor, epilepsy, autism, intracranial arachnoid cyst, chromosomal abnormalities, cardiovascular disease, and endocrine disorders, and others. As a result, all subjects in our study were previously healthy, experienced only minor head trauma, and did not require any surgical interventions.

Multiple follow-up CT scans were performed in certain cases for medical reasons. However, not all scans were included in the analysis since our primary focus was on establishing normal growth curves rather than correlating growth with head trauma. To ensure appropriate selection of scans, we established the following criteria: (1) For patients under one year of age with multiple scans taken at different months, one CT scan obtained at a different month of age was evaluated for each, (2) for patients over one year of age with multiple scans taken at different years, one CT scan obtained at different year of age was evaluated for each, and (3) if multiple scans were obtained at the same age in months or years, we evaluated only the scan least affected by head injury.

### Data collection

We conducted a retrospective collection of CT scans and evaluated various medical variables pertaining to the patients, including sex, age, modified child’s Glasgow coma scales (CGCS)[18], and CT findings. To quantitatively assess the intracranial volume (ICV), PCFV, CBMV, and brainstem volume (BSV), the Digital Imaging and Communications in Medicine (DICOM) CT data were imported into MATLAB R2020b (MathWorks, Natick, MA, USA). The target areas were manually segmented using the image segmenter app in MATLAB (https://www.mathworks.com/help/images/ref/imagesegmenter-app.html). We segmented areas of the medulla oblongata and the pones, treating them as BSV. A representative segmentation of ICV, PCFV, CBMV, and BSV is shown in Figure 1. The DICOM slices had a width of 5 mm. By employing these procedures, we were able to calculate the volume (in milliliter, mL) related to the above parameters. It is worth noting that this method has been utilized in our previous studies [19–21].

**Fig. 1.**
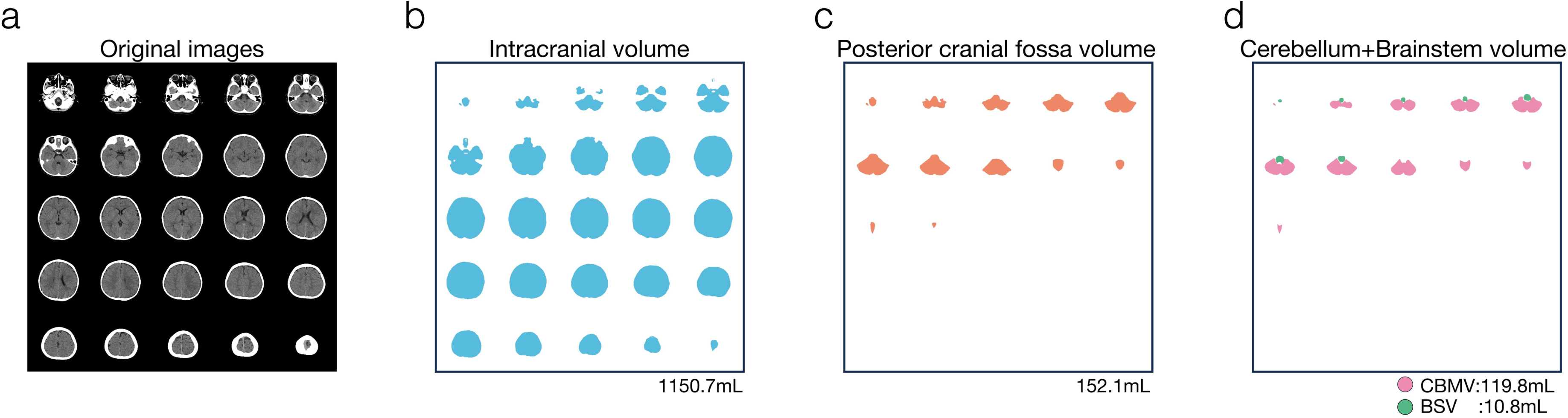
Segmentation of CT images. Typical CT images of a child are shown in (a). The segmentation of the intracranial volume is displayed, with a calculated volume of 1150.7mL in (b). The segmentation of the posterior cranial fossa volume is displayed, with a calculated volume of 152.1mL in (c). The segmentation of the cerebellum volume (CBMV) and brainstem volume (BSV) is displayed, with calculated volume of 119.8mL and 10.8mL, respectively, in (d).

To assess the relationship among intracranial structure, we calculated two types of ratios. The first ratio was PCFV/ICV (PCFV divided by ICV), and the second was (CBMV + BSV)/PCFV (the sum of CBMV and BSV divided by PCFV). We considered the combined volume of CBMV and BSV as the volume of the main structures in the posterior cranial fossa.

### Statistical analyses

Categorical data were presented as frequencies (percentages). For continuous variables with a normal distribution, mean ± standard deviation (SD) was reported, while for variables with a non-normal distribution, the median with 25th-75th quartiles was used. Spearman correlation coefficients were calculated to assess the relationship among the parameters.

To account for the uneven distribution of age, we examined the distribution and defined age-related clusters for adjustment purposes. Mean values and 95% confidence intervals (CIs) were calculated for the parameters. Differences between age-related clusters or genders were evaluated by assessing overlapping or non-overlapping CIs. Non-overlapping CIs were considered indicative of statistical significance, while overlapping CIs suggested a lack of statistical significance [22]. Additionally, unpaired T-tests were employed to compare the age-related clusters, and Bonferroni correction was applied to account for multiple comparisons. Statistical significance between clusters was determined at corrected p-values of <0.05.

The measurements of each parameter were plotted according to months after birth, and best-fit logarithmic curves were generated. Statistical analyses were conducted using the Statistical and Machine Learning Toolbox of MATLAB R2020b.

### Data availability statement

The data used in this study are available from the corresponding authors upon reasonable request and after additional ethics approval.

## Results

### Baseline characteristics

A total of 277 patients were initially enrolled, but 90 patients were subsequently excluded. Among the remaining 187 patients, one outlier aged over 11 years was excluded, resulting in 234 CT scans from 186 patients under 10 years of age being included for evaluation (Fig.2). Out of these CT scans, 97 (41.5%) were obtained from female patients, and the median time interval after birth was 12 months (range: 5-44 months). Based on our criteria for selecting multiple CT scans, twice CT scans were obtained in 40 patients (17 females, 42.5%), and three times in four patients (1 female, 25.0%). Notably, a substantial portion of the study population (49.1%) consisted of children under one year of age, with a total of 115 cases.

**Fig. 2.**
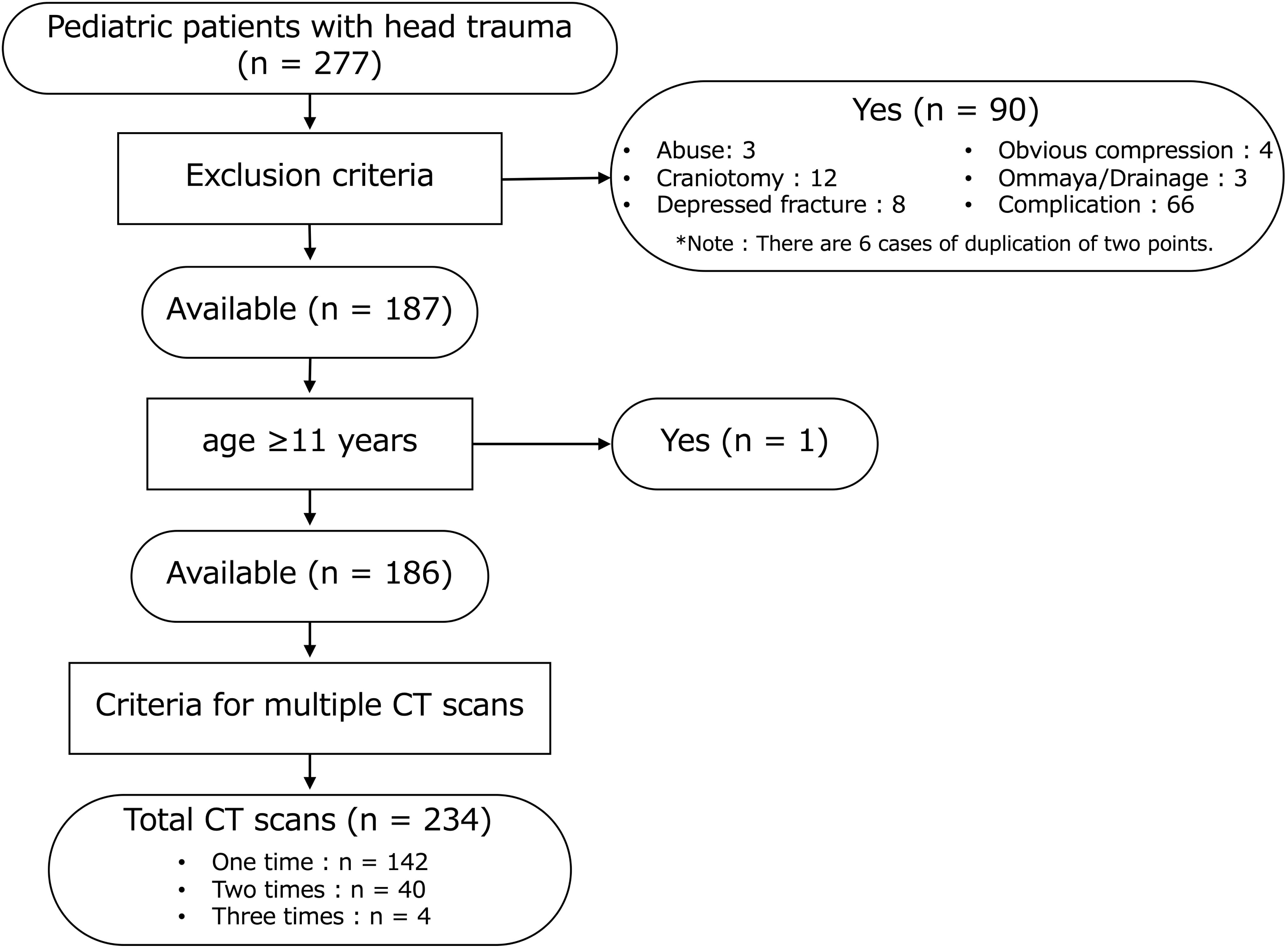
Patients enrollment and CT scans investigated in the study. The flow chart depicts the inclusion and exclusion criteria for pediatric patients with head trauma who were included in this study. Initially, 277 patients were enrolled, but 90 patients were subsequently excluded. One patient who was over eleven years old was excluded from the study, leaving the remaining participants for analysis. Consequently, a total of 186 patients were included in the study. Based on our criteria for selecting multiple CT scans, a total of 234 CT scans were obtained for analysis.

The majority of CT scans had a CGCS value of 15, observed in 199 scans (85.0%). CGCS values of 14 were found in 28 scans (12.0%), and values between 9 and 13 were observed in 7 scans (2.9%). Fractures were present in 109 scans (46.6%), including 101 linear fractures (92.7%) and 8 depressed fractures (7.3%). Intracranial bleeding was observed in 68 scans (29.1%), consisting of 34 acute subdural hematomas (50.0%), 27 acute epidural hematomas (39.7%), 14 subarachnoid hemorrhages (20.6%), 3 intracerebral hematomas (4.4%), and 2 intraventricular hemorrhages (2.9%). There were 6 cases of duplication data points and 3 cases of triplicated data points.

### Correlation analysis

Significant positive correlations were observed among four parameters, namely ICV, PCFV, CBMV, and BSV (Table 1). Particularly noteworthy is the strong correlation between PCFV and CBMV, with a correlation coefficient of 0.99, which was statistically significant (*p* = 7.73 × 10^−233^).

**Table 1.** Correlation coefficients between parameters.

| **Combinations between parameters** | **Correlation coefficient** | ***p*-value** |
| --- | --- | --- |
| ICV – PCFV | 0.96 | $6.99 \times 10^{-130}$ |
| ICV – CBMV | 0.95 | $1.50 \times 10^{-115}$ |
| ICV – BSV | 0.93 | $3.88 \times 10^{-100}$ |
| PCFV – CBMV | 0.99 | $7.73 \times 10^{-233}$ |
| PCFV – BSV | 0.95 | $4.13 \times 10^{-119}$ |
| CBMV – BSV | 0.94 | $3.08 \times 10^{-110}$ |

BSV, brainstem volume; CBMV, cerebellum volume; ICV, intracranial volume; PCFV, posterior cranial fossa volume.

### Age-related clusters

To address the uneven distribution of the study population across different age groups, we categorized the CT scans into fifteen age-related clusters. These clusters include: 0 month, 1 month, 2-3 months, 4-5 months, 6-7 months, 8-9 months, 10-11 months, 1 year-1 (age range 12 to 17 months), 1 year-2 (age range 18 to 23 months), 2 years, 3 years, 4 year, 5-6 years, 7-8 years, and 9-10 years. Table 2 presents the distribution of CT scans performed within each cluster for both males and females. It is important to note that the population distribution between males and females was uneven due to a higher frequency of CT scans for head trauma in males compared to female.

**Table 2.** Number of CT scans among age-related clusters

| Age-related clusters | Male | Female | Total |
| --- | --- | --- | --- |
| 0m | 5 | 3 | 8 |
| 1m | 8 | 8 | 16 |
| 2-3m | 11 | 4 | 15 |
| 4-5m | 10 | 10 | 20 |
| 6-7m | 18 | 6 | 24 |
| 8-9m | 13 | 11 | 24 |
| 10-11m | 4 | 4 | 8 |
| 1y-1 | 11 | 11 | 22 |
| 1y-2 | 5 | 7 | 12 |
| 2y | 6 | 7 | 13 |
| 3y | 8 | 8 | 16 |
| 4y | 10 | 6 | 16 |
| 5-6y | 10 | 3 | 13 |
| 7-8y | 9 | 7 | 16 |
| 9-10y | 9 | 2 | 11 |
| Total | 137 (58.5%) | 97 (41.5%) | 234 |

The table presents the number of CT scans distributed among age-related clusters. The cluster labeled as "1y-1" corresponds to an age range of 12 to 17 months, while the cluster labeled as "1y-2" corresponds to an age range of 18 to 23 months.
m, month; y, year.

### Growth curves

The mean values of each age-related cluster, calculated from the total number of CT scans, were plotted sequentially for ICV (Fig.3a), PCFV (Fig.3b), CBMV (Fig.3c), and BSV (Fig.3d). The growth curves for all four parameters consistently increased with each age-related cluster, and a stabilization of the plots was observed after 4 years of age. Furthermore, the error bars representing the 95% CIs showed minimal overlap among the clusters in all four plots until 4 years of age. Detailed figures can be found in Supplementary Table 1.

**Fig. 3.**
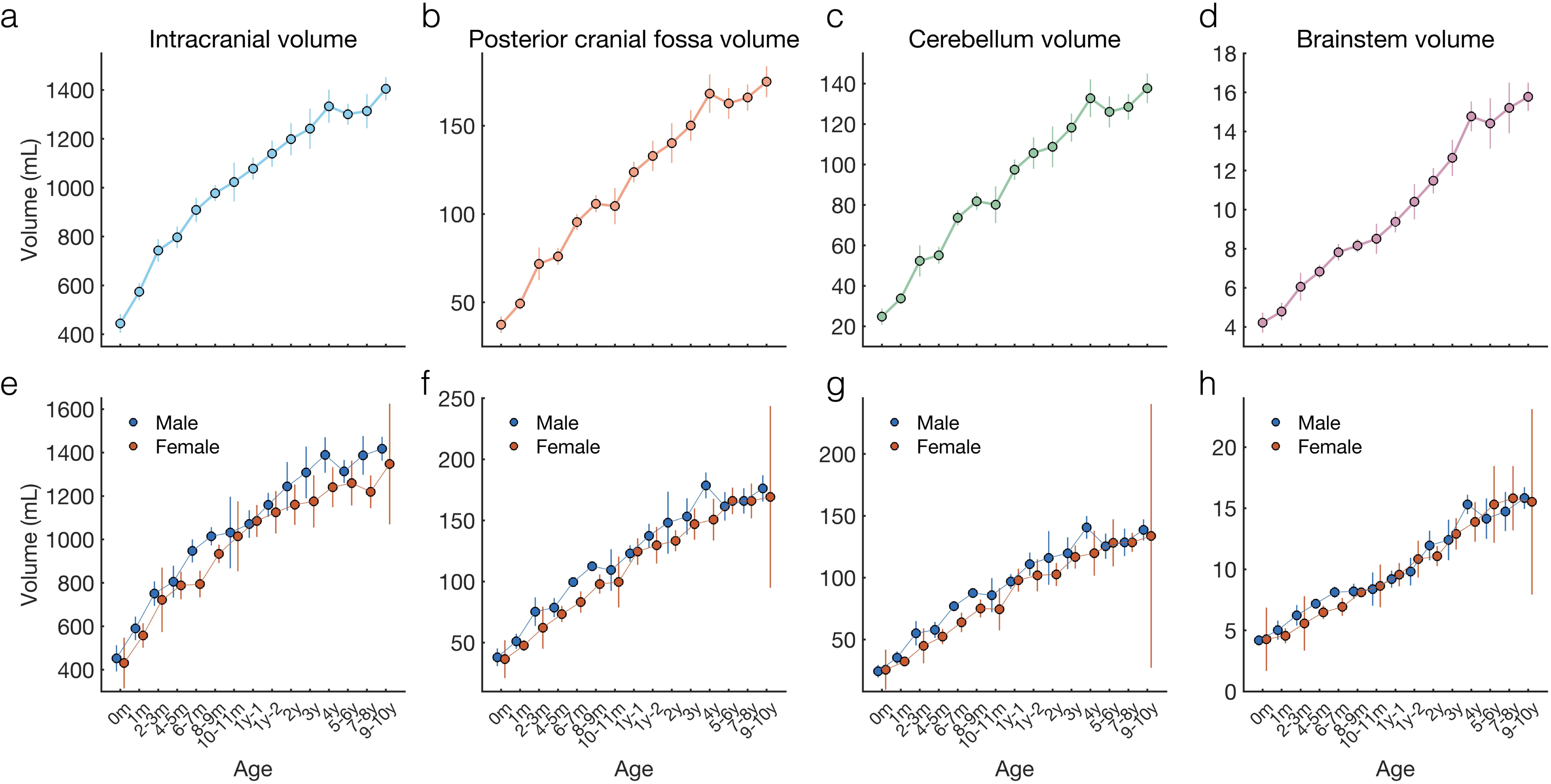
Growth curves for ICV, PCFV, CBMV, and BSV. The mean values, calculated in each age-related cluster, are sequentially plotted for intracranial volume (ICV), posterior cranial fossa volume (PCFV), cerebellum volume (CBMV), and brainstem volume (BSV). The panels in the upper row (a, b, c, and d) represent the results obtained from total CT scans, while the panels in the lower row (e, f, g, and h) display the results obtained from CT scans in males and females. Error bars indicate the 95% confidence intervals.

In the lower row (Fig.3e, 3f, 3g, and 3h), plots generated separately for males and females are shown. With the except of the 6-7 months and 8-9 months clusters, the error bars for males and females exhibited overlapping in each cluster. It is important to note that, due to the small sample size of females, the 95% CIs indicated a wide range, particularly in the 9-10 years cluster where there were only two females. Therefore, in this study, we concluded that we were unable to observe noticeable differences between males and females due to the small sample size. Consequently, in the subsequent analyses, we presented results obtained from the total number of CT scans.

### Statistical differences among each age-related cluster

To evaluate the differences in volumes between two different age-related clusters for each parameter, unpaired T-tests were conducted. To account for multiple comparisons, the Bonferroni correction was applied by multiplying the obtained p-values by 105, which considered the total number of combinations (_15_C_2_). The volume differences with corrected p-values less than 0.05 were presented as a color-scaled matrix (Fig.4). Notably, significant differences in all four parameters were observed between two different age-related clusters up to approximately two years of age (Fig.4a, 4b, 4c, and 4d).

**Fig. 4.**
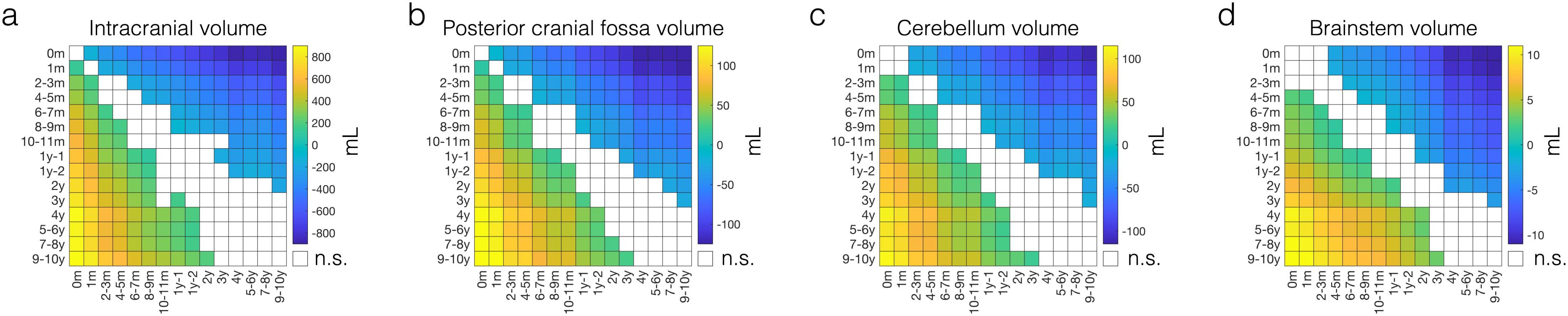
Color-scaled matrix of significant volume differences. The matrix displays differences among age-related clusters in intracranial volume (a), posterior cranial fossa volume (b), cerebellum volume (c), and brainstem volume (d). Significant positive differences, indicating larger volumes in the longitudinal axis compared to the horizontal axis, are presented by shades ranging from green to yellow. Significantly negative differences, indicating smaller volumes in the longitudinal axis compared to the horizontal axis, are presented by shades of blue. Only differences that reached statistical significance with corrected p-values <0.05 using the Bonferroni method are color-coded. n.s., non-significant.

### Changes in ratios

We present the ratios calculated by dividing PCFV by ICV (PCFV/ICV) (Fig.5a). Additionally, we present the ratios calculated by dividing the sum of CBMV and BSV by PCFV ((CBMV + BSV)/PCFV) (Fig.5b). These ratios exhibit dynamic changes throughout the developmental process and show an increasing trend after birth. In the PCFV/ICV ratio, we observed a continuous increase until approximately 4 years of age, after which it stabilized at a consistent value. In the (CBMV+BSV)/PCFV ratio, we observed an increasing trend until the age range of 12-17 months (1y-1), beyond which it also reached a consistent level.

**Fig. 5.**
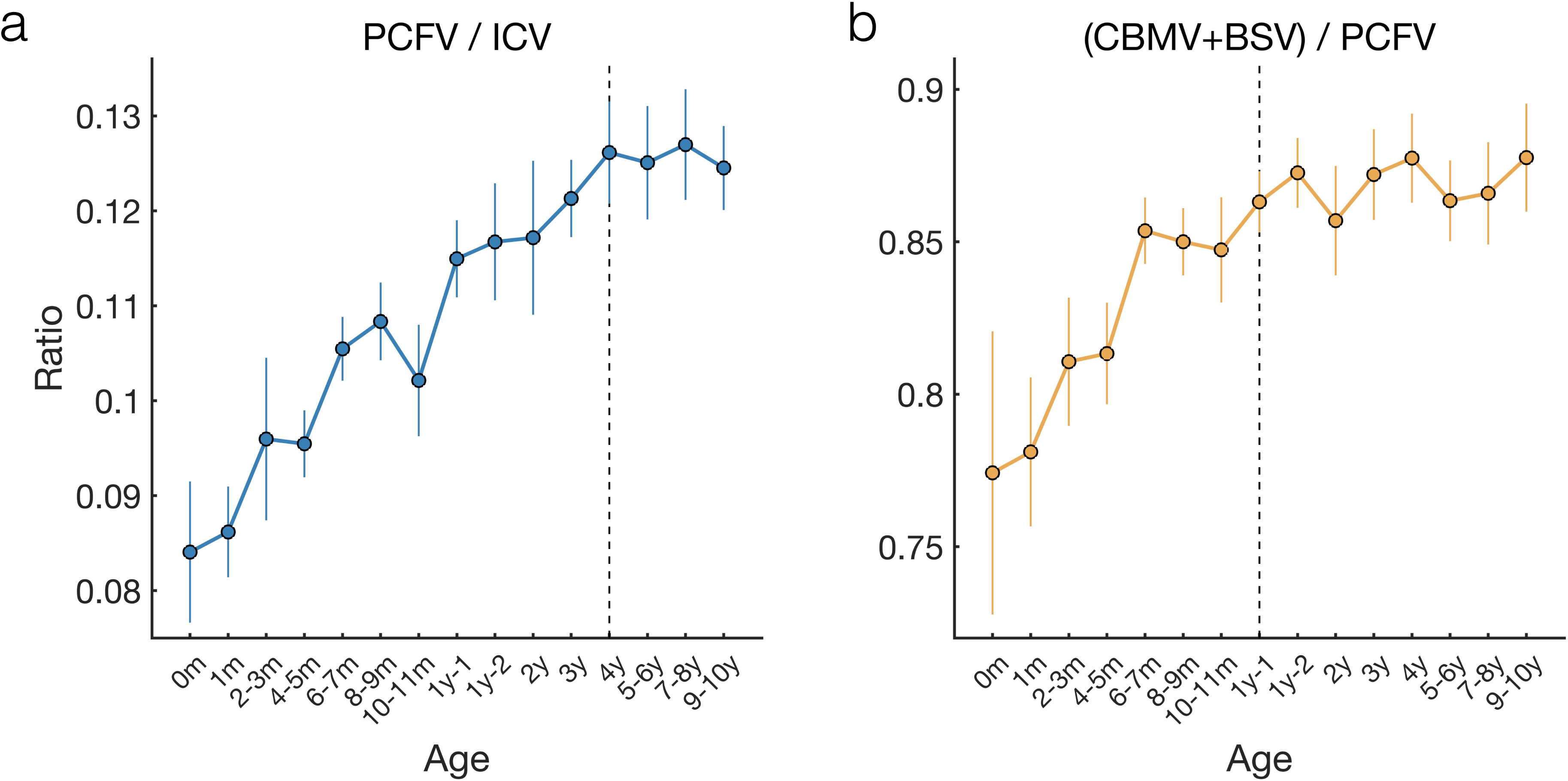
Ratios among intracranial structures. The ratios obtained by dividing the posterior cranial fossa volume (PCFV) by the intracranial volume (ICV) are sequentially plotted across age-related clusters (a). The x-axis represents the age-related clusters ranging from 0 month to 9-10 years. The rations obtained by dividing the sum of the cerebellum volume (CBMV) and brainstem volume (BSV) by PCFV are displayed (b). 1y-1 corresponds to the age of 12-17 months, and 1y-2 corresponds to the age range of 18-23 months. m, month; y, year

### Best-fit logarithmic curves

Best-fit logarithmic curves were generated by scatter plots against age in months. The equations of the curves were as follows: y = 198.04log(x+1) + 495.94 for ICV (Fig.6a), y = 31.14log(x+1) + 32.73 for PCFV (Fig.6b), y = 25.53log(x+1) + 21.21 for CBMV (Fig.6c), and y = 2.73log(x+1) + 2.52 for BSV (Fig.6d). For all four parameters, the growth rate was faster during the early months and gradually slowed down after approximately 20 months of age.

**Fig. 6.**
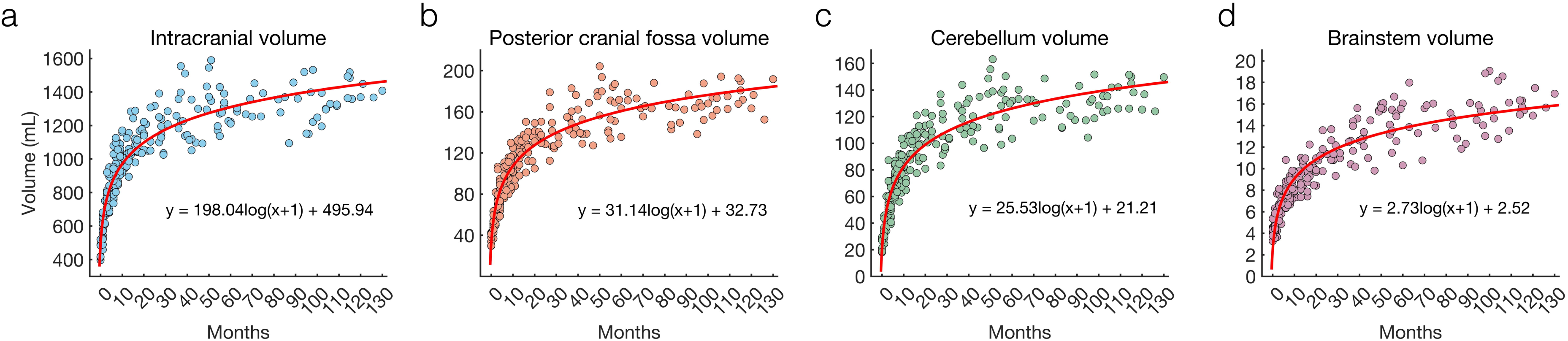
Best-fit logarithmic curves. Scatter plots against age in months are shown for intracranial volume (a), posterior cranial fossa volume (b), cerebellum volume (c), and brainstem volume (d). The best-fit logarithmic curves are indicated as red curves.

## Discussion

In this study, we generated normal growth curves for ICV, PCFV, CBMV, and BSV using DICOM images of CT scans obtained from Japanese pediatric patients with minor head trauma. A total 234 CT scans were included, with approximately half of the cases (115 out of 234) involving patients under one year of age. Positive correlations were observed among the investigated parameters, with the strongest correlation observed between PCFV and CBMV. The growth curves for all four parameters exhibited similar patterns, characterized by a two-phase process. This process involved rapid growth until approximately 4 years of age, followed by a stabilization phase. Furthermore, we examined the ratio of PCFV to ICV and the sum of CBMV and BSV to PCFV. The PCFV/ICV ratio demonstrated continuous increases until approximately 4 years of age, while the (CBMV+BSV)/PCFV ratio showed an increasing trend until one year of age, after which it reached a consistent level.

A two-phase process, characterized by rapid growth of PCFV from birth to approximately 3 to 4 years of age, followed by a stabilization phase, has been previously reported [12, 13]. In our study, we confirmed this growth pattern not only for PCFV but also for ICV, CBMV, and BSV. It is natural for these parameters to exhibit the same growth pattern given the positive correlation among them. While previous studies have reported a positive correlation between PCFV and the volume of the infratentorial structures [12, 23], we provided new insights by demonstrating that PCFV exhibits the strongest positive correlation with CBMV. Given the similar growth patterns observed in all four parameters, we initially expected that ratios such as PCFV/ICV and (CBMV + BSV)/PCFV would remain constant from birth. However, our findings revealed an increasing trend in these ratios from birth onwards. In the case of PCFV/ICV, the proportion of PCFV gradually increased until 4 years of age, after which it stabilized. Similarly, in the (CBMV + BSV)/PCFV ratio, the proportion of the sum of CBMV and BSV continued to gradually increase until one years of age, followed by stabilization. These findings suggest that the proportion of infratentorial space at birth is relatively smaller compared to subsequent stages of development. Additionally, at birth, the extra-axial space of the posterior cranial fossa appears to be relatively larger compared to subsequent stages of development. It is worth noting that not only the growth curves of volumes but also the growth curves of ratios demonstrate a two-phase process.

The previous studies indicated differences in PCFV between males and females, with females having statistically smaller PCFV [12, 13]. However, our study did not find significant differences between males and females. It is important to note that our sample size, particularly for females, may have been inadequate to detect gender-related differences. The proportion of females in our study was approximately 40%, which could be lower than the proportion of males due to the enrollment criteria focused on children with minor head trauma. In a previous study examining head injuries in individuals aged 15 to 79 years, the female population accounted for only 23% [24]. These findings suggest that both pediatric and adult populations have a higher incidence of head trauma in males compared to females.

Previous studies have utilized either CT [12, 13, 25] or magnetic resonance imaging (MRI) [2, 5, 10, 11, 23, 26] to calculate the volume of the posterior cranial fossa. Both CT and MRI have been considered equally valid for volume calculation. In this study, we chose to utilize CT scans to create normal growth curves. Previous research using CT scans reported that PCFV reaches approximately 150mL by 3 to 4 years of age [13], which is consistent with our findings. Although we used CT scans with a 5mm slice width, which raised concerns about lower resolution, a previous CT study that employed a slice width of 2mm or less reported pediatric normal growth curves for intracranial volume [27], similar to our results. These consistencies between previous findings and our study outcomes validate the methodology and outcomes of our study.

In this study, our assumption was that minor head trauma would have no significant impact on the growth of intracranial structures. To minimize the influence of head trauma, we established specific criteria for selecting CT scans. While post-traumatic hydrocephalus can develop after a brain injury, with an incidence ranging from 0.7 to 50% [28, 29], the effect of head trauma on the volume of infratentorial structures remains unknown. However, our main objective in this study was to determine normal growth curves for the infratentorial structures, rather than establishing a correlation between growth and head trauma. Further research is necessary to investigate this relationship. It is worth noting that a previous study utilized CT scans with head trauma to generate normal growth curves for intracranial volume [14]. Nevertheless, it is important to acknowledge that the inclusion of patients with head trauma is a significant limitation of this study.

Our study has several additional limitations. Firstly, we were unable to collect data on low-birth-weight infants, which could have potentially influenced the growth curves presented in our study. Secondly, it is important to note that our study only included data from Japanese children, and there is a possibility that racial differences may have had an impact on our results. However, considering that our findings were consistent with those from other populations [12, 13], we believe that these racial differences can be considered negligible Thirdly, while one advantage of our study was the inclusion of a significant number of patients under one year of age, we acknowledge that the sample size for the 0m and 10-11m age-related clusters were relatively small. Lastly, it is worth noting that our study only provided results for children up to 10 years of age, and the growth curves for children older than 10 years remain unknown.

## Conclusions

Our study conducted on Japanese pediatric patients with minor head trauma provides important insights into the normal growth curves of posterior cranial fossa structures. We have demonstrated a two-phase process characterized by rapid growth and stabilization phases, not only in the growth curves for volumes but also in the changes observed in the ratios of PCFV/ICV and (CBMV + BSV)/PCFV. Comparing values obtained from groups with different mean and SD values poses a challenge. To overcome this, we propose the use of z-normalization, which involves subtracting the mean values and dividing by SD. For instance, direct comparison between PCFV values obtained at birth and one year of age would be invalid, as PCFV doubles during that period. By employing z-normalization, it is possible to accurately compare and analyze data from different age groups. We believe that assessing the volume of the posterior cranial fossa structures in pediatric patients with Chiari malformation while considering these growth-related variations holds the potential to provide new insights into the relationship between the posterior cranial fossa structures and Chiari malformation.

## Compliances with Ethical standards

### Ethics approval

The Ethics Committee of Osaka Women’s and Children’s Hospital (Izumi, Japan, approval no. 1568) provided ethical approval for this study, which was conducted in accordance with the Declaration of Helsinki guidelines for experiments involving humans.

### Informed consent

Informed consent was obtained using the opt-out method from our center’s website because of the retrospective and noninvasive nature of the study.

### Funding

The Japan Society for the Promotion of Science (JSPS) KAKENHI [JP21K16629 (Hiroaki Hashimoto)] supported this work.

## Data Availability

Data availability statement
The data used in this study are available from the corresponding authors upon reasonable request and after additional ethics approval.

## Abbreviation

BSV: brainstem volume
CBMV: cerebellum volume
CGCS: modified child’s Glasgow coma scales
CI: confidence intervals
CT: computed tomography
DICOM: Digital Imaging and Communications in Medicine
ICV: intracranial volume
m: month
mL: milliliter
MMC: myelomeningoceles
MRI: magnetic resonance imaging
PCFV: posterior cranial fossa volume
SD: standard deviation
y: year

