## Supplemental Table for "Growth patterns and ratios of posterior cranial fossa structures in the Japanese pediatric population: A study utilizing CT scans"

Supplemental Table 1

**Supplemental Table 1**

| Intracranial volume (mL) | 0m | 1m | 2-3m | 4-5m | 6-7m | 8-9m | 10-11m | 1y-1 | 1y-2 | 2y | 3y | 4y | 5-6y | 7-8y | 9-10y |
|---|---|---|---|---|---|---|---|---|---|---|---|---|---|---|---|
| Mean | 444.35 | 574.18 | 742.97 | 796.78 | 908.90 | 977.50 | 1023.18 | 1078.05 | 1139.16 | 1198.68 | 1241.70 | 1333.24 | 1300.65 | 1313.63 | 1404.68 |
| SD | 45.84 | 66.19 | 83.74 | 93.89 | 116.85 | 77.11 | 95.16 | 100.02 | 84.02 | 108.42 | 154.36 | 126.21 | 70.18 | 131.18 | 70.36 |

| Posterior cranial fossa volume (mL) | 0m | 1m | 2-3m | 4-5m | 6-7m | 8-9m | 10-11m | 1y-1 | 1y-2 | 2y | 3y | 4y | 5-6y | 7-8y | 9-10y |
|---|---|---|---|---|---|---|---|---|---|---|---|---|---|---|---|
| Mean | 37.39 | 49.33 | 71.79 | 76.00 | 95.45 | 105.81 | 104.50 | 123.75 | 132.90 | 140.17 | 150.11 | 168.17 | 162.59 | 165.99 | 174.93 |
| SD | 5.58 | 6.53 | 16.62 | 10.19 | 10.66 | 11.45 | 12.21 | 13.16 | 13.46 | 18.62 | 16.33 | 20.40 | 14.43 | 13.86 | 13.05 |

| Cerebellum volume (mL) | 0m | 1m | 2-3m | 4-5m | 6-7m | 8-9m | 10-11m | 1y-1 | 1y-2 | 2y | 3y | 4y | 5-6y | 7-8y | 9-10y |
|---|---|---|---|---|---|---|---|---|---|---|---|---|---|---|---|
| Mean | 24.79 | 33.77 | 52.34 | 55.10 | 73.68 | 81.83 | 80.14 | 97.44 | 105.66 | 108.76 | 118.21 | 132.77 | 126.03 | 128.53 | 137.69 |
| SD | 4.74 | 5.22 | 13.92 | 8.98 | 8.94 | 10.31 | 10.87 | 11.31 | 12.20 | 16.65 | 13.06 | 17.49 | 12.77 | 11.79 | 10.80 |

| Brainstem volume (mL) | 0m | 1m | 2-3m | 4-5m | 6-7m | 8-9m | 10-11m | 1y-1 | 1y-2 | 2y | 3y | 4y | 5-6y | 7-8y | 9-10y |
|---|---|---|---|---|---|---|---|---|---|---|---|---|---|---|---|
| Mean | 4.22 | 4.79 | 6.06 | 6.83 | 7.82 | 8.16 | 8.51 | 9.38 | 10.41 | 11.48 | 12.65 | 14.77 | 14.41 | 15.21 | 15.77 |
| SD | 0.62 | 0.85 | 1.28 | 0.74 | 1.00 | 0.81 | 0.92 | 1.23 | 1.42 | 1.07 | 1.73 | 1.43 | 2.13 | 2.42 | 1.07 |

The mean and standard deviation (SD) values calculated for each age-related cluster are presented. The phase labeled as “1y-1” corresponds to an age range of 12 to 17 months, while the phase labeled as “1y-2” corresponds to an age range of 18 to 23 months. Figure 3a, 3b, 3c, and 3d were created using these mean values.

m, month; y, year.
